# The impact of deconfliction on humanitarian access and healthcare delivery in Gaza: a qualitative study of international healthcare workers

**DOI:** 10.64898/2026.07.29.26359296

**Authors:** Eli Lesher, Cole Petersen, James Smith, Shatha Elnakib

## Abstract

**Background:** Deconfliction is a central pillar of humanitarian risk mitigation in contemporary armed conflicts. Intended as a notification system through which parties to a conflict can mitigate against harm to humanitarian personnel and infrastructure, its effectiveness is dependent on respect for humanitarian protections. Since October 2023, repeated Israeli attacks on deconflicted sites and routes in Gaza, alongside increasing restrictions on humanitarian movement as notification-based coordination was replaced by a permission-based system, have raised fundamental questions about the utility and function of deconfliction. This study examines how deconfliction shaped humanitarian access, healthcare delivery, and Palestinian health infrastructure from the perspective of international healthcare workers in Gaza since 2023.

**Methods:** We conducted semi-structured interviews with 18 international healthcare providers who worked in Gaza between January 2024 and December 2025. Participants completed 31 deployments across 16 healthcare facilities. Interviews were analyzed using inductive thematic analysis.

**Results:** Two major themes emerged. First, participants described deconfliction as an unstable and ineffective means of ensuring humanitarian protection or access. They reported delays, denials, sudden revocations of movement permissions, and strikes on deconflicted sites. These conditions disrupted clinical care, delayed transfers and evacuations, limited facility access, and exposed both patients and providers to ongoing risk. Second, participants described deconfliction as a mechanism of control that shifted responsibility for protection from parties to a conflict to humanitarian actors and redirected resources and personnel away from Palestinian hospitals toward NGO (Non-Governmental Organization) facilities, eroding Palestinian healthcare autonomy.

**Conclusion:** Rather than a neutral protection mechanism, deconfliction operated as a conditional authorization regime through which Israel regulated humanitarian movement, medical supply chains, and healthcare delivery. This mechanism rendered protection contingent on compliance, allowing Israel to reframe attacks as humanitarian operational failures rather than violations of international law by military actors. Deconfliction also facilitated the substitution of Palestinian healthcare with internationally managed structures that were dependent on Israeli approval. By undermining local health system autonomy, deconfliction was instrumentalized within a broader Israeli strategy to deliberately inflict conditions to bring about the physical destruction of life in Gaza. These findings raise urgent questions about humanitarian actors’ obligations when operating within non-neutral systems and underscore the need to prioritize direct support for local healthcare institutions.

## Introduction

Over the course of more than two and a half years, Israel’s sustained military assault on Gaza and against the Palestinian people has rendered the occupied territory one of the deadliest places on Earth, including for people afforded explicit protection under international law: among them, civilians, humanitarian workers, and healthcare personnel. As of January 2025, it is estimated that at least 75,200 people had been killed by direct violence alone, a figure that continued to rise in the subsequent months [1]. As of April 2026, at least 593 aid workers, 145 civil defense workers, and more than 1,700 healthcare workers have been killed in Gaza [2]. The targeting of civilians and civilian infrastructure not only caused mass death, injury, and disability but also incapacitated those tasked with mitigating the effects of Israel’s genocide. The systematic erosion of humanitarian capacity raises several urgent questions: why has it become so dangerous to deliver aid and care in Gaza, and what has become of the mechanisms that supposedly exist to prevent such harm?

One such mechanism is deconfliction, which has formed a central pillar of humanitarian risk mitigation strategies in Gaza and other armed conflict contexts. Adapted from military practice into humanitarian operations in the early 2000s, deconfliction refers to a notification-based system intended to reduce the risk of attacks on aid workers, medical facilities, and humanitarian convoys by informing parties to a conflict of the locations and movements of protected actors [3]. In practice, deconfliction requires humanitarian organizations to share detailed information—geographic coordinates, movement routes, precise timings, and personnel data— through a neutral third-party intermediary, typically the United Nations (UN), which then communicates with relevant military actors [4]. Efforts to formalize deconfliction took shape following the 2011 North Atlantic Treaty Organization (NATO) intervention in Libya. These efforts included the development of a structured Humanitarian Notification System for Deconfliction (HNS4D) as well as guidance that followed United Nations (UN) Security Council resolution 2286 in 2016, which promoted “enhanced information exchanges and real-time coordination with medical and humanitarian actors” as a technocratic tool to facilitate compliance with legal obligations and increase accountability when attacks occur [5, 6, 7]. Critically, deconfliction neither changes existing protections guaranteed under international law nor does it generate new legal protections. Rather, deconfliction was intended as a supplementary notification system to help parties to a conflict mitigate against accidental harm to protected actors. The logic of deconfliction rests on the premise that combatants are incentivized to avoid direct attacks on humanitarian or healthcare personnel and infrastructure, since such attacks may incur legal, diplomatic, political, and other costs [8]. Existing literature, however, suggests that the effectiveness of deconfliction remains highly contingent on the willingness of conflict parties to respect humanitarian protections [9]. Deconfliction mechanisms have been implemented in Afghanistan, Iraq, Libya, South Sudan, Syria, Ukraine, and during Israel’s past violent escalations in Gaza in 2008-2009 and 2014 [5]. In some contexts, such as Ukraine, notification systems have facilitated humanitarian movements with relatively few reported incidents [10]. In others, most notably Syria, repeated attacks on deconflicted hospitals and other protected sites prompted concerns that sharing information may expose humanitarian actors to greater risk when belligerents seek to target protected sites [11]. Scholars and humanitarian organizations have therefore questioned whether deconfliction inadvertently substitutes legal protections for limited contractual arrangements dependent on military cooperation [12].

Since October 7th, 2023, violence and extremely limited humanitarian access in Gaza have again drawn deconfliction processes into the spotlight. Deconfliction arrangements used during prior Israeli military assaults in Gaza relied on voluntary information-sharing systems coordinated through United Nations Office for the Coordination of Humanitarian Affairs (UNOCHA), which then communicated information to the Israeli Coordination and Liaison Administration (CLA) [5]. Beginning in late 2023, however, a newly implemented system required humanitarian organizations to coordinate directly with the CLA—a unit of the Israeli military—which operated a model based on permission rather than notification [13].

Available UNOCHA operational data suggest that, as humanitarian movement became increasingly centralized through Israeli-controlled approval systems, denials and obstructions to planned humanitarian activities rose from 31.4% between January and early May 2024 to 51.6% between mid May 2024 and December 2025 [Compiled from 14]. At the same time, repeated attacks on deconflicted sites and routes—including the April 2024 World Central Kitchen (WCK) convoy strike that killed seven aid workers travelling on a pre-notified route—have intensified scrutiny of the efficacy and purpose of the current deconfliction mechanism in Gaza [15, 16]. Subsequent attacks affecting hundreds of humanitarian personnel, including the March 2025 attack on a Palestine Red Crescent Society (PRCS) convoy that killed 15 aid workers, have further undermined confidence in deconfliction as a protection mechanism, raising fundamental questions about its utility [17, 18].

International healthcare workers who worked across public hospitals, NGO facilities, and temporary field hospitals throughout Gaza have observed how the deconfliction mechanism functioned in practice and how it affected the delivery of care. Existing discussions on deconfliction have largely focused on legal and policy analyses, while comparatively little attention has been paid to how frontline healthcare workers experience these systems and how they affect the ability to deliver humanitarian and health services. Drawing on qualitative interviews with international healthcare providers who worked in Gaza between 2024 and 2025, this study examines how deconfliction shaped humanitarian access, healthcare delivery, and Palestinian health infrastructure.

## Methods

### Study design

This study employed semi-structured interviews to explore the experiences of international healthcare workers who have worked in Gaza since October 2023. Because the study aimed to examine how humanitarian coordination and access systems shaped clinical care and hospital operations, sampling intentionally focused on frontline healthcare workers directly involved in patient care. International staff were selected as the deconfliction mechanism disproportionately served international organizations, and as such, they were more likely to have frequent interactions with it than local healthcare staff.

### Sampling and participants

Purposive and snowball sampling strategies were employed, from which 18 participants were recruited, with recruitment continuing until the study team agreed that thematic saturation had been reached. Initial participants were identified through collaboration with an NGO involved in the coordination of international medical teams to work in Gaza, with snowball sampling used thereafter. Eligible participants had to be an international healthcare worker who had worked in Gaza on at least one occasion between January 2024 and December 2025.

A degree of professional diversity in the sample was ensured by purposively including participants with a range of clinical roles, including physicians (n=11), nurses (n=6), and nurse practitioners (n=2). The sample included 11 male and 8 female participants. Clinical specialties represented in the sample included pulmonology, emergency medicine, obstetric and regional anesthesiology, internal medicine, pediatric critical care, lactation consulting, pediatrics, nephrology, surgery (orthopedic, general, and trauma), primary care, critical care, and wound care. Participants were primarily based in the United States and Jordan.

Participants collectively completed 31 deployments within the study period, completing either one (n=11), two (n=5), three (n=2), or four deployments (n=1). These deployments spanned 16 distinct healthcare facilities, including public hospitals (n=8), NGO-operated hospitals (n=2), and temporary NGO-run facilities (n=6). Overall, 72% of deployments occurred in public hospitals, 7% in NGO hospitals, and 21% in temporary NGO facilities.

### Data Collection

Data were collected through semi-structured interviews conducted between May 2025 and February 2026 via videoconferencing software or telephone, and lasted approximately one hour. All interviews were conducted in English. Interviews were recorded and transcribed.

### Data Analysis

Transcripts were analyzed using an inductive thematic approach informed by Braun and Clarke’s 2006 framework [19]. Following familiarization with the transcripts, the researchers generated initial codes that reflected recurring concepts and patterns. These codes were then grouped into potential themes and were iteratively refined through discussion among the research team to ensure that they accurately represented the data.

Two members of the research team independently reviewed and coded transcripts to enhance reliability. Reliability was assessed from a selection of transcripts, with discrepancies resolved through discussion until a consensus was reached. While participants discussed many topics due to the semi-structured format, this paper focuses only on themes that emerged in relation to deconfliction and associated consequences for healthcare delivery.

### Ethical Considerations

Ethical approval was sought from the Johns Hopkins University Homewood Institutional Review Board, for which exemption was granted (Ref: HIRB00021096, dated 10 April 2025). Verbal informed consent was obtained from all participants before each interview. All data were de-identified to preserve participant confidentiality.

## Results

Two interrelated themes emerged from participants’ accounts of deconfliction in Gaza. First, participants described deconfliction as failing to ensure humanitarian protection and access, characterizing it as an unstable and unreliable mechanism that not only fell short of its intended purpose but also reshaped how care could be delivered. Second, participants viewed deconfliction as a mechanism of control that inverted the responsibility for protection and contributed to the fragmentation of the Palestinian healthcare system. Together, these themes illustrate how deconfliction functioned as both a failed protection mechanism and as an active force that manipulated humanitarian practice and eroded Palestinian healthcare autonomy.

### 1. Deconfliction Failed to Ensure Humanitarian Protection and Access

#### A. Delays, denials, and revocations

Participants described the deconfliction process as having two components: (1) petitioning the Coordination and Liaison Administration (CLA) to designate areas as “safe zones” and (2) informing Coordination of Government Activities in the Territories (COGAT) of movements prior to traveling to hospitals. Yet participants frequently described how permissions could be revoked at any stage, routes altered without notice, access to facilities denied, and requests rejected.

> “There’s some days where… you’re trying to go to your work site… and you just get turned down, and they don’t have to give you any reason.”
>
> “The IDF (Israeli Defense Forces) via the CLA can always… be like … you’re going to lose your deconflicted status, because we have… operations planned for this area”

Even when approval was granted, it did not necessarily translate into access, and long wait times were commonly reported.

> “We waited hours and hours… literally asking Israeli forces to stop bombing so that we can cross… you can smell gunpowder and burning flesh.”
>
> “One time… it was a six-hour wait to just get a green light [approval to move]… and then they may not give you a green light… you have to go home.”

#### B. Failure of protection in deconflicted spaces and routes

Participants noted that even when approvals were secured for their movements, deconflicted hospitals, safe houses, and travel routes were still exposed to bombardment and gunfire, with healthcare workers sustaining injuries.

> “Even when we were in our safe zone, we didn’t feel safe… the green zone next to us was being bombed continuously.”
>
> “European [Gaza] Hospital was shaking the entire time I was there from explosions. Some of the… other people in my group were thrown into a wall. One had his tooth broken, and one had his finger broken from explosions. The windows were blown out on the third floor of the hospital.”

Participants described repeated bombings and attacks on and near their deconflicted routes after receiving permission to travel, including attacks that interrupted and endangered convoys or that necessitated rerouting.

> “Israel was dropping bombs near our safe house, and then… Israel dropped the bomb… a couple blocks away from us as we were driving to the hospital.”
>
> “We were in our car, and the grenade hit… launched from one of the military boats from the ocean… it hit… maybe 50 meters… from us to see and hear the detonation… and that was in the safe zone… you couldn’t be too sure of your own safety.”
>
> “Not even… 500 yards away, there would be a smoking building that was bombed… So there was a lot of… last-minute route changes.”

Participants also described learning about the loss of so-called “safe zone” status outside of formal coordination channels, including from forced evacuation leaflets dropped by Israeli warplanes.

> “We started out in a safe house… then they flew over, and they dropped these flyers… And the flyers just have a picture of the city and an arrow, and it’s like, we’re going to bomb this place, move… so our safe house was no longer safe, and so we ended up… living in the hospital.”

Several participants emphasized that the spatial boundaries of “safe zones” were functionally meaningless.

> “The TSP (Trauma Stabilization Point)… was fenced by… a mesh. So… if there was something to happen… you would be very, very much exposed to it. So we were housed … in the so-called safe zone, and also the TSP… was… within the safe zone, but it’s… clear that you can’t count on that, and we’ve experienced… firings coming quite close to our TSP… a bullet … hit into the tent next to us where one of our… colleagues was taking a nap. So he was missed by the bullet by like a meter… that happened… about three meters or four meters from our tent”

#### C. Clinical consequences of the deconfliction process

Participants recounted how delays and denials resulting from deconfliction interfered with international medical teams’ ability to operate, and described how dependence on permission to move made humanitarian organizations ineffective. Many providers expressed frustration, helplessness, and distress arising from their inability to deliver medical care as a result of the deconfliction process.

> “There were many days where they would tell us, No, you can’t leave… delay it an hour. There was… a lot of frustration… because several of the days we weren’t able to get to the hospital at the time we had originally planned, it was cutting into… our patient care time.”
>
> “It’s genuinely exhausting… to be… a trained medical professional… a few kilometers away from where… you’re supposed to be working, and … not be able to go there because … an outside agency … decided on that day you can’t go do your job. That’s pretty tough.” “The grand scheme… it almost felt designed to limit any true change.”

Participants described how the deconfliction process shaped risk calculations and humanitarian obligations, sometimes forcing teams to choose between adherence to deconfliction procedures and patient survival. Participants shared how performing complex and time-sensitive procedures amid ongoing bombardment fundamentally compromised the provision of care.

> “I was doing a very critical procedure on a patient in the emergency department… a pericardiocentesis, where you withdraw blood from around the heart… You don’t want to puncture the heart muscle, because you can kill them… your hand has to be steady… as soon as I got the needle in, a loud explosion right outside the window… my ears were ringing… So there was nothing safe about it.”

A participant recalled their experience trying to deploy an ambulance to rescue a newborn who went into cardiac arrest.

> “They had to go into the red zone… and the ambulance driver was like, “We are ready. You let us know.”… The Palestinian doctor that had to accompany the patient was like, “I’m ready to go. We will take the risk.” And the parents said the same thing… We were like, we don’t want anybody to go at risk, but… this kid’s gonna die if we don’t send them… we didn’t really have a leg to stand on as internationals. The national team all said, “If we’re willing to take the risk, we’re going to take the risk.”

Another shared how repeated movement cancellations by Israel impacted care for pediatric patients who were scheduled for medical evacuation.

> “The mission was about medical evacuation from the north to the south of mostly pediatric patients who were dying… things would be set up, and then Israel would decide - ‘Nope, it’s canceled’ again and again and again and again. So a lot of these organizations, even with the best intentions… became pretty ineffective.”

One provider described how the deconfliction mechanism prohibited them from accessing hospitals during periods of the highest clinical need.

> “There was a third [foreign surgeon whose] NGO… mandated that they leave the hospital and go back to their safe house every day, which means that he wasn’t there for 90% of the trauma cases because they came in at night.”

In sum, participants described deconfliction as unstable, unpredictable, incapable of ensuring physical safety, and detrimental to their ability to provide adequate medical care.

### 2. Deconfliction as a Mechanism of Control and Health System Fragmentation

#### A. Surveillance and intimidation

Rather than enhancing the safety of humanitarian personnel and protected infrastructure, participants often described that deconfliction increased their visibility to Israeli military forces, while questioning whether the system fulfilled its stated purpose of reducing risk to humanitarian and medical personnel. Many pointed to repeated warnings from the United Nations that, despite compliance with extensive coordination and notification procedures, deconfliction would not guarantee safety.

> “There’s definitely no semblance of safety. And the UN makes that very, very clear… that, yes, you are deconflicted, and yes, the Israeli authority knows where you are, but that’s not a guarantee of safety.”
>
> “When we were in Cairo… the United Nations and the WHO (World Health Organization), they told us… don’t think for a moment that… you are safe. Despite all this communication and everything, we cannot guarantee your safety at any moment, you could be targeted.”
>
> “Yeah, so the deconflicting—it’s all a sham.”

Participants described numerous encounters with drones and tanks used by the Israeli military to intimidate them while travelling on pre-approved routes. Several participants reported that on occasion they considered not informing the CLA of their movements so as to avoid harassment and the risk associated with disclosing their movements.

> “There was a DJI (Da-Jiang Innovations) Mavic drone right next to me, and he said, do not pull out your phone. Just keep on looking straight. And that [drone] just kept on circulating around us… that drone stayed there for a few minutes.”
>
> “They have all our information, the vehicles’ description, people’s description. I feel like that was… a way to scare us that… the drone is there, the tanks are there, and… they come in with… guns close to you.”
>
> “Nobody trusted it… some people even suggested that it’s better not to tell them about movements… but people were scared.”

Several described their expectation that healthcare workers would be protected from attack, but stated that they lost confidence in this assumption after experiencing the situation in Gaza.

Participants reported feeling that their status as healthcare workers did not provide protection and instead made them more visible and vulnerable.

> “I felt this kind of fake, artificial sense of safety because I thought… healthcare workers have this level of immunity, even in war zones… they shouldn’t be targeted. But when I got there… I realized that’s not true… In fact, it’s the opposite. Healthcare workers are specifically targeted and killed.”

#### B. Deconfliction as redistribution of responsibility and risk

The deconfliction system was also described as having shifted the onus for humanitarian safety away from combatants, insofar as the deconfliction process elevated adherence to coordination protocols as a prerequisite for protection. This was said to shift the burden of risk management onto humanitarian and medical teams, who were forced to decide whether to delay or cancel humanitarian movements, or to proceed without full clearance.

> “It’s an interesting situation because… in most… conflict zones, the burden of responsibility for keeping humanitarian workers safe is… on the combatants… In Gaza… early on in the conflict, if we were driving to a job site and there was an incident… the whole international community would be up in arms about it, because everyone kind of assumes… the combatants are responsible for keeping humanitarians safe. Whereas now that everyone bought into the system… if something were to happen and we didn’t follow the rules, now they can very clearly say… we have rules in place that weren’t followed.”
>
> “It seemed, early on, like maybe a good idea that we… follow the processes and… coordinate and notify our movements, but in reality just kind of restricted everyone. And it took that responsibility and shifted it to us instead of… them. And… by the time that anyone… really realized… you can’t go back on it, right? Once the system’s in place, you can’t be like, “You know what? We’re not going to coordinate movements anymore, because we think it’s your job. It’s your job to keep us safe.” You just can’t, you can’t go back on that.”

Several participants invoked the World Central Kitchen incident, noting that the attack had a chilling effect on humanitarian actors and reinforced the perception that adherence to deconfliction protocols did not ensure safety.

> “They were telling us this is a known convoy… deconflicted… and it was completely obliterated… we had just taken that route the day before.”
>
> “That incident… showed that… there was still quite a palpable chance this could happen again to our team.”

#### C. Channeling personnel and supplies away from Palestinian hospitals

Participants perceived the deconfliction system as shaping where international staff, medical supplies, and operational capacity could be deployed. In their accounts, the system concentrated resources and relative safety within NGO-managed facilities while Palestinian hospitals remained under-resourced and vulnerable to attack, despite being formally deconflicted.

> “Working in the field hospitals… my impression is that they tend to be fairly safe. I think it’s different if you’re a medical provider working in a Ministry of Health hospital or… a Palestinian facility.”
>
> “A consistent pattern all along is that… the Ministry of Health… hospitals, do get targeted… The… facilities that the NGOs have set up do not, and I’m not aware of any incidents… at those facilities since the start of the conflict.”

Participants stated that Israeli military attacks on public hospitals affected where international organizations were willing to work. Many NGOs diverted staff to the few public hospitals that were perceived as safer.

> “One of the problems with European [Gaza] Hospital from the standpoint of internationals is that… it was actually the only [public] hospital that hadn’t been attacked in Gaza at that point… Every NGO wanted to send people there.”

When those hospitals were also attacked, NGOs eventually diverted international staff and resources away from public hospitals altogether, even if this meant fragmenting Palestinian health system capacity.

> “The… field hospital had not been struck by any bombs, so it was considered to be safe… I can’t stress enough… this hospital is a field hospital… it was not meant to be a major, tertiary care hospital.”

Many healthcare workers also criticized the deconfliction process as a way to limit critical supplies reaching Palestinian institutions. Participants described how their movements were used as a method to restrict supplies and to funnel them away from Palestinian hospitals in particular. They explained that Israeli control over supply movements consistently privileged field hospitals and undermined the ability of Palestinian hospitals to provide adequate care.

> “I’ll ask for… vancomycin… Israel will allow some of that stuff to enter through international groups, but then not take them to local Palestinian hospitals… Because [the] majority of people will go to local hospitals… and therefore that’s where most of them will die, because there’s nothing. But the field hospital will have a little bit more. So it’s like a drop… in the middle of nothing.”
>
> “The system of coordinating movements… allows the IDF to… keep tabs on where everybody is… and funnel where medical resources go, and restrict their access.”
>
> “There’s nothing safe about it… this is all a tactic to delay things and prevent things from getting in and out.”
>
> “[The field hospital] had some equipment better than us. So if we had a really, really hopeless case… we’ll send them to [a] field hospital… it’s only local hospitals that are left with nothing.”

Taken together, participants described deconfliction not only as a system governing humanitarian movement but also as one that shaped the distribution of health personnel, supplies, and clinical capacity, impeding the ability to provide care to critically ill and injured patients. In their accounts, international healthcare workers described that coordination requirements and movement restrictions concentrated resources within internationally managed facilities while limiting support for Palestinian hospitals.

## Discussion

This study suggests that deconfliction and humanitarian coordination mechanisms in Gaza have functioned not as neutral systems for effective civilian and humanitarian protection, but as conditional authorization regimes that regulate the humanitarian system by titrating when, where, and by what means humanitarian and health services can be delivered. Participants consistently described deconfliction as an unstable and ineffective means to ensure safety, despite extensive coordination and surveillance requirements. Rather than reducing risk, deconfliction was frequently understood as a process that increased visibility to the Israeli military, while simultaneously functioning to restrict essential movements and delay or prevent the provision of care. Importantly, participants emphasized that United Nations personnel explicitly communicated that deconfliction did not guarantee safety, underscoring the apparent powerlessness of internationally mandated institutions to meaningfully constrain violence and repeated violations of humanitarian protection and access. In this way, a system purportedly designed to enable the safe delivery of essential humanitarian services has functioned to regulate the conditions under which care can be administered. The deployment of seemingly neutral humanitarian and procedural language when promoting the deconfliction process has allowed Israel to imply that humanitarian access has been facilitated and that international obligations have been upheld. Yet, behind this veneer lies an attempt to limit—if not entirely strangulate— humanitarian access.

Our findings also show how the current deconfliction process in Gaza inverts the protective principles underpinning international humanitarian law; deconfliction mechanisms were intended as supplementary and voluntary notification systems through which parties to a conflict can minimize the risk of harm to protected actors. Yet participants described a permission-based system in which compliance with coordination shifted responsibility for protection away from combatants and onto humanitarian personnel. The requirement to repeatedly request authorization for movement transformed humanitarian protection from a legal entitlement into a conditional and revocable privilege. Participants described being forced to choose between delaying or forgoing care to maintain procedural compliance, or proceeding without authorization, and thereby accepting heightened risk. Under Israel’s deconfliction system, attacks on humanitarian and health personnel could be reframed as failures of organizational compliance rather than failures of parties to the conflict to adhere to their legal obligations. Participants described an increasing burden to “prove” the legitimacy of their movements and activities.

Israeli military attacks on several deconflicted locations and movements highlight how Israel has used the system to avert blame. Israeli explanations after the WCK attack, as well as other attacks including the killing of a UN worker in Rafah, shifted blame from the Israeli military and onto humanitarian organizations: claiming that WCK workers had taken the wrong route, even though an Australian government report found that the Israeli military was not aware that the convoy had strayed from the route until the day after the attack, or claiming that specific humanitarian actors did not have deconflicted status in the case of the UN worker killed in Rafah [20, 12, 16].

Furthermore, our findings indicate that the Israeli-controlled system of deconfliction not only affected individual humanitarian movements but also undermined Palestinian health capacity by fragmenting healthcare delivery and facilitating a process of controlled substitution.

Internationally-managed facilities were presented as comparatively protected and resourced, while Palestinian hospitals faced greater supply shortages and more frequent attacks, with Israel bombing 94% of Gaza’s public hospitals by May 2024 [21]. Israel’s coordination mechanism was perceived as funneling supplies and capacity away from Palestinian hospitals. At the same time, the overarching “humanitarian space” remained entirely dependent on Israeli authorization for entry and the movement of all staff and supplies. The result was not simply the emergence of parallel infrastructure in response to the decimation of Palestinian health system capacity, but the progressive replacement of the Palestinian health system with an international system whose operations remain entirely contingent on Israeli military approval. In that way, deconfliction ultimately allowed Israel to dictate which institutions functioned and what forms of care were possible. Israel’s repeated attempts to expel international personnel, deny deconfliction for safe movement, ban UNRWA, and deregister tens of humanitarian organizations left Palestinians to navigate a severely incapacitated healthcare system, with an encumbered humanitarian system left to offer partial, temporary relief [22, 23, 24, 25].

Participants described the destruction of Gaza’s health infrastructure as consistent with what Ghassan Abu Sittah refers to as the “biosphere of war,” in which restrictions on medical supplies, the obstruction of evacuation pathways, and the incapacitation of humanitarian operations were central features of Israel’s military strategy rather than incidental byproducts of violence [26]. This distinction is particularly important in the context of genocide, whose material acts under the Genocide Convention include “deliberately inflicting on the group conditions of life calculated to bring about its physical destruction in whole or in part” [27]. Taken together, these actions and the resulting displacement of healthcare delivery away from Palestinian institutions may therefore be understood not only as a failure of humanitarian protection, but as its active manipulation to further the destruction of conditions necessary for the collective survival of the Palestinian people. While deconfliction is commonly framed as a technical and politically neutral risk mitigation tool, our findings suggest that in Gaza, it is deeply embedded in both the politics and practice of genocide.

Given these findings, a residual question concerns what humanitarian workers and organizations should do when operating under such conditions. Participants consistently described the deconfliction process as fundamentally ineffective and harmful while simultaneously feeling unable to withdraw from it because doing so would further restrict their ability to maintain an operational presence and reach patients, resulting in even greater immediate suffering. Our findings suggest that it is important to deliberate over the feasibility and impact of collective refusal to participate in such harmful systems, even when doing so may reduce immediate operational access, especially since operational presence alone cannot be treated as evidence of humanitarian effectiveness or ethical legitimacy. Participants repeatedly described a sense that humanitarian organizations, in attempting to preserve access, were no longer able to fulfil the humanitarian imperative or meet their medical obligations, not only because they were unable to adequately protect civilians or deliver sufficient care, but because continued participation in authorization-dependent systems risked contributing to the erosion of Palestinian health autonomy.

Indeed, our results raise questions about the political effects of contemporary deconfliction regimes globally and support existing critiques that such systems can shift responsibility for humanitarian protection onto aid organizations while normalizing extensive surveillance and movement control [10, 11]. Because providers explicitly linked these systems to delayed transfers and interrupted evacuations, medication shortages, an inability to provide standard treatment, and preventable patient deaths, we argue that highly restrictive forms of permission-based humanitarian access have normalized a state in which care is increasingly contingent on approval from parties to a conflict.

At a minimum, humanitarian organizations should critically and transparently interrogate coordination arrangements that shape the provision of care through their humanitarian programs. They must also intentionally prioritize direct support for Palestinian healthcare institutions in the face of clear attempts to displace Palestinian institutional autonomy. Participants’ accounts suggest that early decisions to adopt deconfliction procedures later made withdrawal or renegotiation difficult once these systems became embedded in operational practice, foreclosing meaningful opportunities for collective reversal. These findings suggest that humanitarian organizations may benefit from developing clearer thresholds for collective refusal or suspension of participation in coordination arrangements when such systems become dominated by one party to the conflict, in order to prevent the normalization of conditions that undermine independent humanitarian action.

Our study has several limitations. Interviews were conducted exclusively with international medical personnel and therefore do not capture the perspectives of Gazan healthcare workers, who have borne the overwhelming burden of caring for patients and who invariably experienced these systems differently. The exclusion of Gazan healthcare workers reflects both practical and ethical challenges associated with conducting interviews under ongoing conditions of insecurity and displacement. Unlike their local counterparts, international healthcare workers were typically deployed for limited periods, retained the possibility of evacuation, had access to organizational resources, and were not the primary targets of Israel’s ongoing genocide. Consequently, the experiences documented here should not be interpreted as representative of healthcare workers in Gaza as a whole; rather, they reflect the perspectives of a group that, despite these relative privileges, still reported significant challenges associated with deconfliction processes.

Second, this study specifically reflects the perspectives of frontline healthcare workers rather than humanitarian administrators, logisticians, diplomats, or coordination personnel. Participants, therefore, primarily experienced the deconfliction system through its effects on clinical care, movement restrictions, and hospital operations. Their perspective may not fully capture broader institutional, political, or operational considerations.

Finally, as with all qualitative research, our findings draw on the experiences of a purposive sample of frontline healthcare workers and are not intended to be generalizable. Rather, they offer a situated understanding of how deconfliction was experienced and navigated by some of the individuals attempting to operate within this system. Our confidence in these findings is bolstered by the fact that we reached theoretical saturation, with accounts and insights fairly consistent across participants.

## Conclusion

Taken together, our findings challenge the premise that deconfliction in Gaza served to protect health workers, reduce risk, or facilitate humanitarian access. Instead, we contend that deconfliction evolved into a highly restrictive form of pseudo-humanitarian governance through which humanitarian movements, the transfer of medical supplies, and healthcare delivery were administratively regulated, conditioned, and strategically directed by Israel, as a dimension of its wider genocide against the Palestinian people. In practice, this not only stymied humanitarian operations but also shaped the distribution of personnel, resources, and clinical capacity, diverting support away from the Palestinian health system and toward externally managed humanitarian structures, further eroding the autonomy of Gaza’s health system. These findings underscore the need for clearer collective thresholds for engagement with, or refusal of, coordination systems that become instrumentalized and subverted by a party to a conflict, alongside prioritization of direct support for Palestinian healthcare institutions.

## Data Availability

The datasets generated during the study are not publicly available because they consist of transcripts containing potentially identifiable and sensitive participant information, but requests for de-identified data will be considered by the corresponding author on reasonable request and subject to approval by the Johns Hopkins University Homewood Institutional Review Board.

## Acknowledgements

This research would not have been possible without the guidance of Dr. Bayan Abdulhaq of the School for International Training in Amman, Jordan. It also benefited from the support of a humanitarian organization that facilitated connections with returning medical staff from Gaza. The authors are grateful to all healthcare worker participants for their time, trust, and valuable contributions to this study.

## Notes

### Competing Interest Statement

The authors have declared no competing interest.

### Author Declarations

Ethical approval was sought from the Johns Hopkins University Homewood Institutional Review Board and was deemed to be exempt (Ref: HIRB00021096, dated 10 April 2025). Verbal informed consent was obtained from all participants before beginning the interviews, and participants were informed of their right to withdraw from the study at any time without consequence, voluntary participation, and the confidentiality of their identities. All data were anonymized to protect participant confidentiality.

